# Partner loss and suicide among older adults: A nationwide cohort study from Austria

**DOI:** 10.64898/2026.08.02.26359535

**Authors:** Anna Schultz, Erwin Stolz, Emilise Pötz, Christian Jagsch, Carlos Watzka, Annette Erlangsen

## Abstract

**Background:** Limited evidence motivated us to examine risk of suicide after partner loss among older adults in Austria.

**Methods:** All married older adults aged ≥65 years and living in Austria (*N*=1,293,557) were followed during 2014-2023. Risk ratios (aRRs) for partner loss were calculated based on adjusted cumulative incidence functions.

**Results:** A total of 357 suicides occurred among widowed older adults. Incidence rates among widowed and not widowed were 56.9 and 29.4 per 100,000 person-years, respectively. High rates were found among widowed males (170.8/100,000) and widowers aged ≥85 (94.1/100,000). Suicide risk was highest in the first month (aRR, 13.6; 95%-CI: 8.7,22.4) but remained elevated up to five years after partner loss (aRR, 2.5; 95%-CI: 2.1, 2.8).

**Conclusion:** Recently bereaved older adults had elevated risks of suicide suggesting monitoring and psychosocial support may be beneficial when bereaved.

**Keypoints:**

- Older adults who had lost a partner had a twofold higher suicide risk than those who had not.
- Suicide risk peaked during the first month after partner loss but remained elevated for up to five years.
- Risk of suicide was more pronounced among widowed males than females.

## Introduction

Older adults have some of the highest suicide rates worldwide, specifically in Austria[1]. Bereavement, particularly the loss of a spouse or partner, has emerged as an risk factor for suicide in late life[2], but this evidence is based on very few studies. In Denmark, older adults were found to have markedly elevated risks of suicide during the first year of widowhood, in particular for males aged 80 years and above [3]. Similarly, widowed males and females were found to have higher suicide rates than married in a national cohort study from Italy[4]. Regarding timing, a Swiss study[5] found that suicide rates among those aged 60 and above were highest during the first week after losing a partner, especially for men, and remained elevated throughout the first year. Beyond the psychological pain and shock, bereavement may lead to social isolation, less emotional and instrumental support, and loneliness[6–8], all of which have been suggested to link bereavement to suicidality among older adults[7].

Due to global population ageing, a higher number of older adults are likely to experience spousal bereavement in the coming decades. The present study aims to examine suicide risks following partner loss among older adults aged 65 years and older in Austria.

## Methods

In this nationwide retrospective cohort study, we included all married Austrian older adults aged 65 and above between 2014-2022 who were followed until 31^st^ of December 2023. Individual-level data from multiple registers (Central Residence Register, Central Civil Status Register, and Educational Attainment Register) were linked through a unique identification number by the Austrian Micro Data Centre (AMDC) at Statistics Austria.

The Central Civil Status Register provided information on date and cause of death, civil status of the deceased, the surviving partner’s identification number, and the surviving partner’s sex. We were able to identify legally married couples and those living in registered partnerships. The date of partner loss was identified through record linkage using the surviving partner’s identification number and date of death. Bereavement information was only available when the deceased partner was aged 65 years or older. Consequently, deaths of partners younger than 65 years were not captured. Linkage accuracy was verified by cross-checking the surviving partner’s sex and confirming that the deceased was recorded as married at time of death.

Suicide deaths were identified via the Central Civil Status Register, which records causes of death according to the 10^th^ revision of the International Classification of Diseases (ICD-10), as X60-84, Y87.0. In line with previous research[5], we excluded dyadic suicides (*n* = 33), defined as couples who died by suicide on the same date using the same method or within 10 days following a homicide. We differentiated between the following suicide methods: poisoning (X60-69), hanging (X70), drowning (X71), firearms (X72-X74), jumping from height (X80), and other (X75-79, X81-84, Y87.0).

In order to account for confounding, we included the following covariates: age (65-74/75-84/≥85 years) and sex (males/females), educational attainment (low/medium/high), country of birth (Austria: no/yes), and area of residence (city/town/rural)[9]. Educational level was classified according to the International Standard Classification of Education (ISCED-1997) and categorised as low (level 1-2), medium (level 3-4) and high (level 5-8).

All individuals were followed from the index date. For newly widowed, the date of partner loss served as the index date. For the remaining older adults who did not experience partner loss during follow-up, an index date was randomly selected from the pool of index dates of newly widowed persons, which had the same year of study entry. This procedure ensured that both groups were followed for a similar length of time. Included older adults were followed until date of suicide, other cause of death, or end of follow-up (31^st^ of December, 2023), whichever occurred first. We further censored cases of migration and divorce. Because exact dates were unavailable, migration was censored at the earliest possible date (1^st^ November), and divorce at the interval midpoint (30^th^ April) in each year.

We calculated overall and sex-, education- and age-specific suicide incidence rates per 100,000 person-years for individuals with and without partner loss. Supported by prior evidence[3–5], we hypothesized that the risk of suicide would be highest immediately after a loss of partner. We estimated stratified and adjusted cumulative incidence functions for suicide among individuals with and without partner loss using proportional hazard survival regression models and G-computation[10]. These adjusted cumulative incidence functions accounted for competing risks (i.e., deaths from other causes) and confounding factors (i.e., age, education, nationality, and area of residence). Based on these estimates, we calculated adjusted relative risk ratios (aRRs) at 1 month, 1 year, 3 years, and 5 years as well as adjusted absolute risk differences at 5 years. To account for potential under-reporting of suicides, we included deaths due to self-harm with undetermined intent (ICD-10: Y10-Y34, Y87.2) in a sensitivity analysis.

Analyses were performed using R 4.4.3[11] and the R-package *riskRegression* (2026.3.11)[12,13], *adjustedCurves* (0.11.3)[10], and *ggplot2* (3.5.1)[14].

## Results

A total of 1,293,577 married older adults aged 65 years or older were included. Of these, 206,136 (15.9%) lost their partner during the 5 years of follow-up. The median follow-up time among those who lost their partner was 2.78 years (IQR=3.58) and 2.58 (IQR=3.91) among those who did not. Out of a total of 1,232 suicide deaths, 357 (29.0%) had lost their partner during the follow-up.

The suicide rate of older adults who lost their partner was 56.9 (95%-CI=51.1, 63.1) per 100,000 person-years, while the rate was 29.4 (95%-CI=27.5, 31.4) among those who had not lost a partner (Table 1). After a partner loss, the highest rates were found for males (170.8, 95%-CI=151.4, 192.0) and those aged 85 years and above (94.1, 95%-CI=77.5, 113.2). Among those who died by suicide after a loss of partner, 14.6% died within the first month after the loss, 23.3% within the first three months, and 47.3% within the first year. Most suicides among older adults who lost their partner occurred by hanging (40.3%), followed by firearms (29.1%), jumping from height (10.4%), poisoning (9.0%), and drowning (2.5%). A comparable distribution of suicide methods was found for older adults who had not lost a partner.

**Table 1:** Suicides, person-years and incidence rates with 95% confidence intervals for individuals with and without partner loss.

|  | Partner loss |  |  | No partner loss |  |  |
| --- | --- | --- | --- | --- | --- | --- |
|  | *n* | Person-years | IR (95% CI) | *n* | Person-years | IR (95% CI) |
| Total | 357 | 627,531 | 56.9 (51.1, 63.1) | 875 | 2,978,133 | 29.4 (27.5, 31.4) |
| Sex |  |  |  |  |  |  |
| Males | 281 | 164,482 | 170.8 (151.4, 192.0) | 756 | 1,720,626 | 43.9 (40.9, 47.2) |
| Females | 76 | 463,049 | 16.4 (12.9, 20.5) | 119 | 1,257,507 | 9.5 (7.8, 11.3) |
| Age |  |  |  |  |  |  |
| 65-74 | 73 | 196,990 | 37.1 (29.0, 46.6) | 383 | 1,906,513 | 20.1 (18.1, 22.2) |
| 74-85 | 172 | 311,535 | 55.2 (47.3, 64.1) | 389 | 930,268 | 41.8 (37.8, 46.2) |
| ≥85 | 112 | 119,006 | 94.1 (77.5, 113.2) | 103 | 141,352 | 72.9 (59.5, 88.4) |
| Education |  |  |  |  |  |  |
| Low | 125 | 276,507 | 45.2 (37.6, 53.9) | 268 | 807,271 | 33.2 (29.3, 37.4) |
| Medium | 214 | 322,863 | 66.3 (57.7, 75.8) | 548 | 1,870,431 | 29.3 (26.9, 31.9) |
| High | 18 | 28,161 | 63.9 (37.9, 101.0) | 59 | 300,432 | 19.6 (14.9, 25.3) |
| Born in Austria |  |  |  |  |  |  |
| Yes | 328 | 562,340 | 58.3 (52.2, 65.0) | 812 | 3,114,287 | 26.1 (24.3, 27.9) |
| No | 29 | 65,191 | 44.5 (29.8, 63.9) | 63 | 491,377 | 12.8 (9.9, 16.4) |
| Urbanization |  |  |  |  |  |  |
| City | 101 | 173,553 | 58.2 (47.4, 70.7) | 178 | 1,009,206 | 17.6 (15.1, 20.4) |
| Town | 112 | 200,187 | 55.9 (46.1, 67.3) | 258 | 1,151,300 | 22.4 (19.8, 25.3) |
| Rural | 144 | 253,790 | 56.7 (47.9, 66.8) | 439 | 1,445,158 | 30.4 (27.6, 33.4) |

*Notes.* IR = incidence rate per 100,000 person-years; CI = confidence interval assuming a Poisson distribution of events.

In stratified and adjusted cause-specific survival analysis, older adults who had lost their partner had a higher suicide risk than those who did not (Supplementary Table 2). The risk was highest during the first month (aRR=13.6; 95%-CI=8.7, 22.4) and declined thereafter, but remained elevated even after 5 years (aRR=2.5; 95%-CI=2.1, 2.8). By the end of follow-up, absolute (adjusted) risk of suicide was 0.31% (95%-CI=0.28, 0.35) for those who lost their partner and 0.13% (95%-CI=0.12, 0.14) for those who did not (Figure 1). In sex-stratified analyses, risk of suicide was more pronounced among widowed males than females (Figure 1 and Supplementary Table 2).

**Figure 1:**
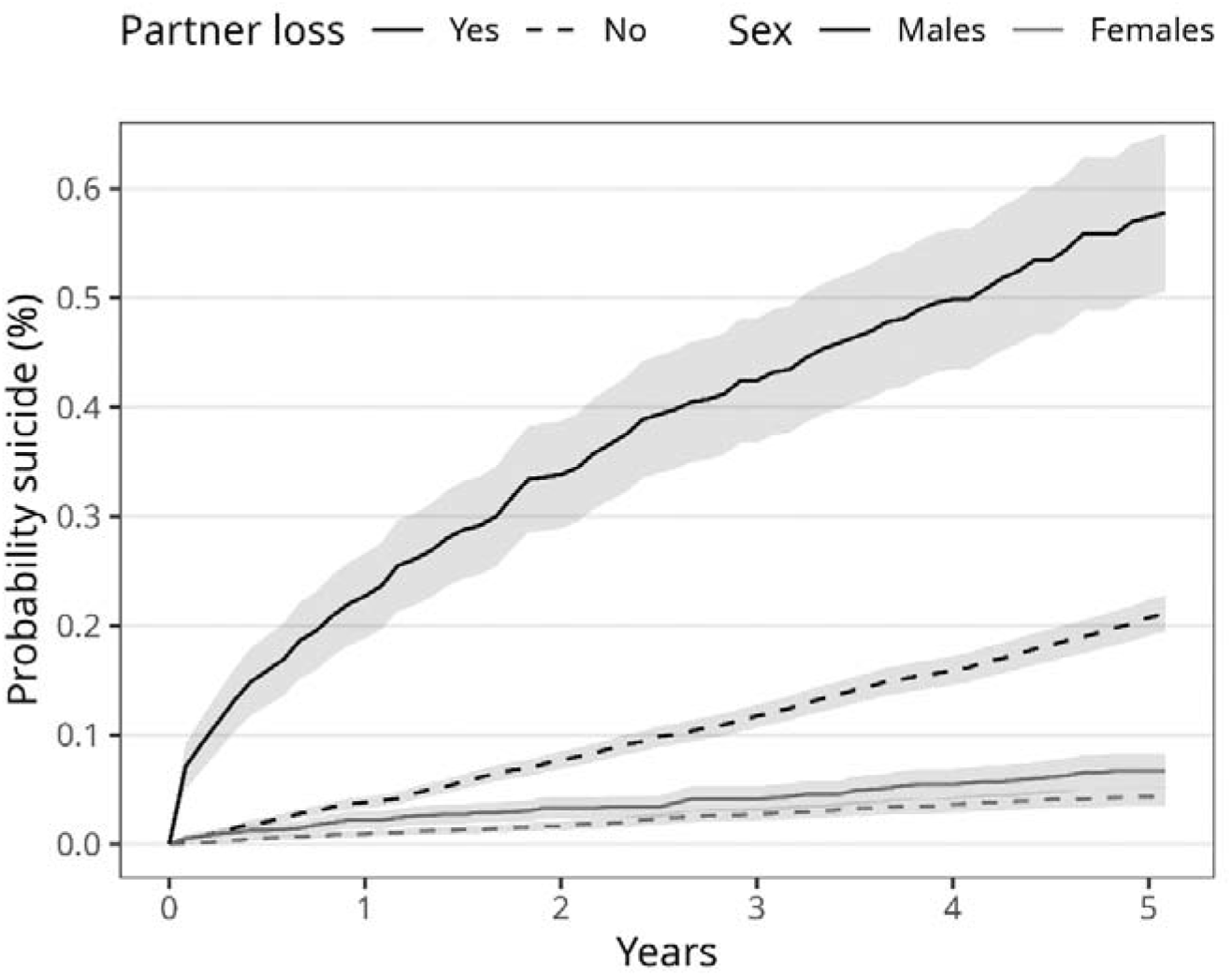
Adjusted probability of suicide stratified by partner loss and sex. Alt text: Line graph depicting the adjusted probability of suicide over five years, stratified by partner loss and sex, showing higher suicide risk among males with partner loss.

When including deaths with undetermined intent in the sensitivity analysis similar patterns were observed (Supplementary Tables 2 and 3, and Supplementary Figure 2).

## Discussion

In this nationwide cohort study, we found considerably higher suicide rates among widowed older adults compared to those who did not lose their partner, specifically among males and those aged 85 years and above. The risk of suicide was highest in the month immediately after the loss of one’s partner, but older adults who had lost a partner also had elevated risks even five years after the loss.

The excess risk among the oldest old is in line with Erlangsen et al.[3] who found that those aged 80 years and above faced the highest suicide risk following spousal loss. As such, our findings align with previous evidence that older males are at an increased risk of suicide following major disruptions in intimate relationships, such as relationship breakdown[15], suggesting that this vulnerability also extends to widowhood. After a partner loss, females seem to receive more support through conversation[16] and increase their social engagement, while men show little change of[17] or decreased social network activities[18].This may leave men less equipped to cope with bereavement and could possibly lead to increased depressive symptoms, loneliness, lower-wellbeing, and poorer adjustment[17–19]. Partner loss has also been linked to declines in physical health and increased frailty[20]. Physical illness has been associated with suicidal ideation among men[21]. More than half of older males who died by suicide were found to have suffered multiple physical health conditions affecting several organ systems[22].

The excess risks of suicide during the first months following the loss of one’s partner, as well as the elevated long-term risk among widowed individuals supports previous findings for older adults[3] and all age groups[23]. Further, we found slightly higher estimates than those reported for older adults aged 60 and above[5].

Our findings highlight the immediate post-loss period as a high-risk period where (clinical) awareness and monitoring is mandated. Interventions addressing complicated grief may reduce symptoms of pathological grief, albeit with limited effect sizes[24]. There seems to be a lack of evidence regarding support for the oldest-old. Also, interventions that address age-specific challenges, such as declining social networks, limited mobility, and increasing frailty, are lacking[24].

Strengths of this study include use of complete, nationwide registry data, and exact dates of partner loss and dying. Limitations include that our analyses were limited to persons in legal partnerships, meaning that cohabiting couples were not included. Second, some of the obtained linkage data were updated on a yearly basis; this implied that the exact dates of divorces were not available, we only had access to information on the year of a divorce.

Third, data on emigration was not available. We assumed that emigration had taken place if information on civil status and area of residence was missing but there was not a record of a death. Fourth, the low autopsy rate in Austria (6.5% of all deaths) implied that some suicide deaths may have gone undetected[25]. Nevertheless, expanding the outcome definition to include events of undetermined intent supported our main findings. Finally, no information on mental disorders or level of psychological distress were available.

In conclusion, older adults who had lost a partner had a twofold higher suicide risk than those who had not. The risk was higher among males (fourfold) than females (twofold) and was greatest immediately after partner loss. Monitoring and psychosocial support for recently bereaved older adults, particularly males, should therefore be considered.

## Data Availability

Access and linkage of register data was approved of and provided by the Austrian Micro Data Center (AMDC) of Statistics Austria following national legislation. The AMDC is a research data infrastructure facility of Statistics Austria that enables research on micro data processed in compliance with data protection regulations. The data used for this research can be accessed by researchers at scientific institutions accredited with the AMDC against a fee.For further information, see https://www.statistik.at/en/services/tools/services/center-for-science/austrian-micro-data-center-amdc.

## Competing interests

The authors declare that they have no competing interests.

## Acknowledgements

This research project was conducted with data from the Austrian Micro Data Centre (AMDC). The AMDC is a research data infrastructure facility of Statistics Austria that enables research on micro data processed in compliance with data protection regulations.

## Declaration of Sources of Funding

This work was supported by the Austrian Academy of Sciences (OeAW) [grant number DATA_2023-08_SAOAA].

## Ethics approval

The study was approved by the Ethics Committee of the Medical University of Graz (EK-number: 1172/2024).

## Notes

### Competing Interest Statement

The authors have declared no competing interest.

## References

1. Davis Weaver N, Bertolacci GJ, Rosenblad E et al. Global, regional, and national burden of suicide, 1990–2021: a systematic analysis for the Global Burden of Disease Study 2021. The Lancet Public Health 2025;10(3):e189–202. 10.1016/S2468-2667(25)00006-4.

2. Beghi M, Butera E, Cerri CG et al. Suicidal behaviour in older age: A systematic review of risk factors associated to suicide attempts and completed suicides. Neuroscience & Biobehavioral Reviews 2021;127:193–211. 10.1016/j.neubiorev.2021.04.011.

3. Erlangsen A. Loss of partner and suicide risks among oldest old: a population-based register study. Age and Ageing 2004;33(4):378–83. 10.1093/ageing/afh128.

4. Grande E, Alicandro G, Vichi M et al. Suicide After Partner’s Death in the Elderly Population: Results From a Nationwide Cohort Study in Italy. The American Journal of Geriatric Psychiatry 2024;32(7):825–31. 10.1016/j.jagp.2024.01.031.

5. Ajdacic-Gross V, Ring M, Gadola E et al. Suicide after bereavement: an overlooked problem. Psychol Med 2008;38(5):673–6. 10.1017/S0033291708002754.

6. Holmes TH, Rahe RH. The social readjustment rating scale. Journal of Psychosomatic Research 1967;11(2):213–8. 10.1016/0022-3999(67)90010-4.

7. Fässberg MM, Orden KAV, Duberstein P et al. A Systematic Review of Social Factors and Suicidal Behavior in Older Adulthood. IJERPH 2012;9(3):722–45. 10.3390/ijerph9030722.

8. Marsa R, Bahmani B, Ebadi A et al. Grief in the elderly: a qualitative content analysis. BMC Geriatr 2025;25(1):540. 10.1186/s12877-025-06027-z.

9. European Commission. Statistical Office of the European Union. Applying the Degree of Urbanisation: A Methodological Manual to Define Cities, Towns and Rural Areas for International ComparisonslJ: 2021 Edition. LU: Publications Office, 2021. https://data.europa.eu/doi/10.2785/706535 (16 Dec. 2025, date last accessed).

10. Denz R, Klaaßen_Mielke R, Timmesfeld N. A comparison of different methods to adjust survival curves for confounders. Statistics in Medicine 2023;42(10):1461–79. 10.1002/sim.9681.

11. R Core Team. R: A Language and Environment for Statistical Computing., version 4.2.3. R Foundation for Statistical Computing, 2023. https://www.R-project.org/.

12. Gerds TA, Kattan MW. Medical Risk Prediction: With Ties to Machine Learning. 1st edn, n.p.: Chapman and Hall/CRC, 2021. 10.1201/9781138384484.

13. Gerds TA, Ohlendorff JS, Ozenne B. riskRegression: Risk Regression Models and Prediction Scores for Survival Analysis with Competing Risks. 2026. 10.32614/CRAN.package.riskRegression.

14. Wickham H. Ggplot2: Elegant Graphics for Data Analysis. Use R! 2nd ed. 2016, Cham: Springer International Publishing_: Imprint: Springer, 2016. 10.1007/978-3-319-24277-4.

15. Wilson MJ, Scott AJ, Pilkington V et al. Suicidality in men following relationship breakdown: A systematic review and meta-analysis of global data. Psychological Bulletin 2025;151(7):819–60. 10.1037/bul0000482.

16. Fischer CS, Beresford L. Changes in Support Networks in Late Middle Age: The Extension of Gender and Educational Differences. The Journals of Gerontology: Series B 2015;70(1):123–31. 10.1093/geronb/gbu057.

17. Niino K, Patapoff MA, Mausbach BT et al. Development of loneliness and social isolation after spousal loss: A systematic review of longitudinal studies on widowhood. J American Geriatrics Society 2025;73(1):253–65. 10.1111/jgs.19156.

18. Yoon H, Park GR, Kim J. Psychosocial trajectories before and after spousal loss: Does gender matter? Social Science & Medicine 2022;294:114701. 10.1016/j.socscimed.2022.114701.

19. Tambellini E, Danielsbacka M, Rotkirch A. Changes in subjective wellbeing during widowhood: Gender differences and the buffering effect of the close social network. JFamRes 2025;37:141–62. 10.20377/jfr-1155.

20. Oberndorfer M, Mogg C, Haider S et al. Partner loss and its effect on frailty trajectories: results from the 13-year follow-up Survey of Health, Ageing and Retirement in Europe (SHARE). J Epidemiol Community Health 2022;76(3):209–15. 10.1136/jech-2021-216637.

21. Jeon GS, Jang SN, Rhee SJ et al. Gender Differences in Correlates of Mental Health Among Elderly Koreans. The Journals of Gerontology Series B: Psychological Sciences and Social Sciences 2007;62(5):S323–9. 10.1093/geronb/62.5.S323.

22. Almeida OP, McCaul K, Hankey GJ et al. Suicide in older men: The health in men cohort study (HIMS). Preventive Medicine 2016;93:33–8. 10.1016/j.ypmed.2016.09.022.

23. Mogensen H, Möller J, Hultin H et al. Death of a Close Relative and the Risk of Suicide in Sweden—A Large Scale Register-Based Case-Crossover Study. PLoS ONE 2016;11(10):e0164274. 10.1371/journal.pone.0164274.

24. Roberts KE, Walsh LE, Saracino RM et al. A Systematic Review of Treatment Options for Grieving Older Adults. Curr Treat Options Psych 2019;6(4):422–49. 10.1007/s40501-019-00191-x.

25. Pritchard C, Hansen L. Examining Undetermined and Accidental Deaths as Source of ‘Under-Reported-Suicide’ by Age and Sex in Twenty Western Countries. Community Ment Health J 2015;51(3):365–76. 10.1007/s10597-014-9810-z.

