## Supplement for "Partner loss and suicide among older adults: A nationwide cohort study from Austria"

Supplementary Table 1: Adjusted risk ratios for partner loss and suicide risk during the first 5 years after loss of partner

|  | Total^a^ | Males^b^ | Females^c^ |
| --- | --- | --- | --- |
|  | aRR (95% CI) | aRR (95% CI) | aRR (95% CI) |
| Follow-up-time |  |  |  |
| 1 month | 13.6 (8.7, 22.4) | 15.7 (9.7, 26.8) | ^d^ |
| 1 year | 5.0 (4.1, 6.1) | 5.9 (4.7, 7.4) | 2.4 (1.4, 3.8) |
| 3 years | 3.1 (2.6, 3.6) | 3.6 (3.1, 4.2) | 1.5 (1.0, 2.1) |
| 5 years | 2.5 (2.1, 2.8) | 2.7 (2.3, 3.2) | 1.5 (1.1, 2.1) |

*Notes*. aRR = adjusted risk ratios, CI = confidence intervals.

^a^ Total: N=1,293,577; events=1,232; C-index=0.707; adjusted for age, education, country of birth, and area of residence.

^b^ Males: N=679,947; events=1,037; C-index=0.609; adjusted for age, education, country of birth, and area of residence.

^c^ Females: N=613,360; events=195; C-index=0.546; adjusted for age, education, country of birth, and area of residence.

^d^ Too few cases for robust estimation.

*Supplementary Table 2: Suicides (including deaths with unclear intent; Y10-Y34, Y87.2), person-years and incidence rates for individuals with and without partner loss*

|  | Partner loss | | | No partner loss | | | |
| --- | --- | --- | --- | --- | --- | --- | --- |
|  | *n* | Person-years | IR (95% CI) | | *n* | Person-years | IR (95% CI) |
| Total | 432 | 627,531 | 68.8 (62.5, 75.6) | | 1,017 | 2,978,133 | 34.1 (32.1, 36.3) |
| Sex |  |  |  | |  |  |  |
| Males | 326 | 164,482 | 198.2 (177.3, 220.9) | | 858 | 1,720,626 | 49.9 (46.6, 53.3) |
| Females | 106 | 463,049 | 22.9 (18.7, 27.7) | | 159 | 1,257,507 | 12.6 (10.8, 14.8) |
| Age |  |  |  | |  |  |  |
| 65-74 | 95 | 196,990 | 48.2 (39, 59) | | 448 | 1,906,513 | 23.5 (21.4, 25.8) |
| 74-85 | 208 | 311,535 | 66.8 (58, 76.5) | | 449 | 930,268 | 48.3 (43.9, 52.9) |
| ≥85 | 129 | 119,006 | 108.4 (90.5, 128.8) | | 120 | 141,352 | 84.9 (70.4, 101.5) |
| Education |  |  |  | |  |  |  |
| Low | 144 | 276,507 | 52.1 (43.9, 61.3) | | 311 | 807,271 | 38.5 (34.4, 43.1) |
| Medium | 261 | 322,863 | 80.8 (71.3, 91.3) | | 633 | 1,870,431 | 33.8 (31.3, 36.6) |
| High | 27 | 28,161 | 95.9 (63.2, 139.5) | | 73 | 300,432 | 24.3 (19, 30.6) |
| Born in Austria |  |  |  | |  |  |  |
| Yes | 397 | 562,340 | 70.6 (63.8, 77.9) | | 948 | 3,114,287 | 30.4 (28.5, 32.4) |
| No | 35 | 65,191 | 53.7 (37.4, 74.7) | | 69 | 491,377 | 14.0 (10.9, 17.8) |
| Area of residence |  |  |  | |  |  |  |
| City | 134 | 173,553 | 77.2 (64.7, 91.4) | | 229 | 1,009,206 | 22.7 (19.8, 25.8) |
| Town | 141 | 200,187 | 70.4 (59.3, 83.1) | | 305 | 1,151,300 | 26.5 (23.6, 29.6) |
| Rural | 157 | 253,790 | 61.9 (52.6, 72.3) | | 483 | 1,445,158 | 33.4 (30.5, 36.5) |

*Notes*. IR = incidence rate per 100,000 person-years; CI = confidence interval assuming a Poisson distribution of events.

*Supplementary Table 3: Adjusted risk ratios for partner loss and suicide risk (including deaths with unclear intent; Y10-Y34, Y87.2)*

|  | Total^a^ | Males^b^ | Females^c^ |
| --- | --- | --- | --- |
|  | aRR (95% CI) | aRR (95% CI) | aRR (95% CI) |
| Follow-up-time |  |  |  |
| 1 month | 14.9 (9.8, 24.3) | 18.2 (11.5, 30.8) | ^d^ |
| 1 year | 4.9 (4.0, 5.8) | 6.0 (4.9, 7.4) | 2.1 (1.3, 3.2) |
| 3 years | 3.1 (2.9, 3.5) | 3.7 (3.1, 4.3) | 1.6 (1.2, 2.1) |
| 5 years | 2.5 (2.2, 2.8) | 2.8 (2.4, 3.2) | 1.6 (1.3, 2.1) |

*Notes*. aRR = adjusted risk ratios, CI = confidence intervals.

^a^ Total: N=1,293,577; events=1,449; C-index=0.690; adjusted for age, education, country of birth, and area of residence

^b^ Males: N=679,947; events=1,184; C-index=0.601; adjusted for age, education, country of birth, and area of residence.

^c^ Females: N=613,360; events= 265; C-index=0.561; adjusted for age, education, country of birth, and area of residence.

^d^ Too few cases for robust estimation.

*Supplementary Figure 1: Adjusted probability of suicide (including deaths with unclear intent; Y10-Y34, Y87.2) stratified by partner and sex*

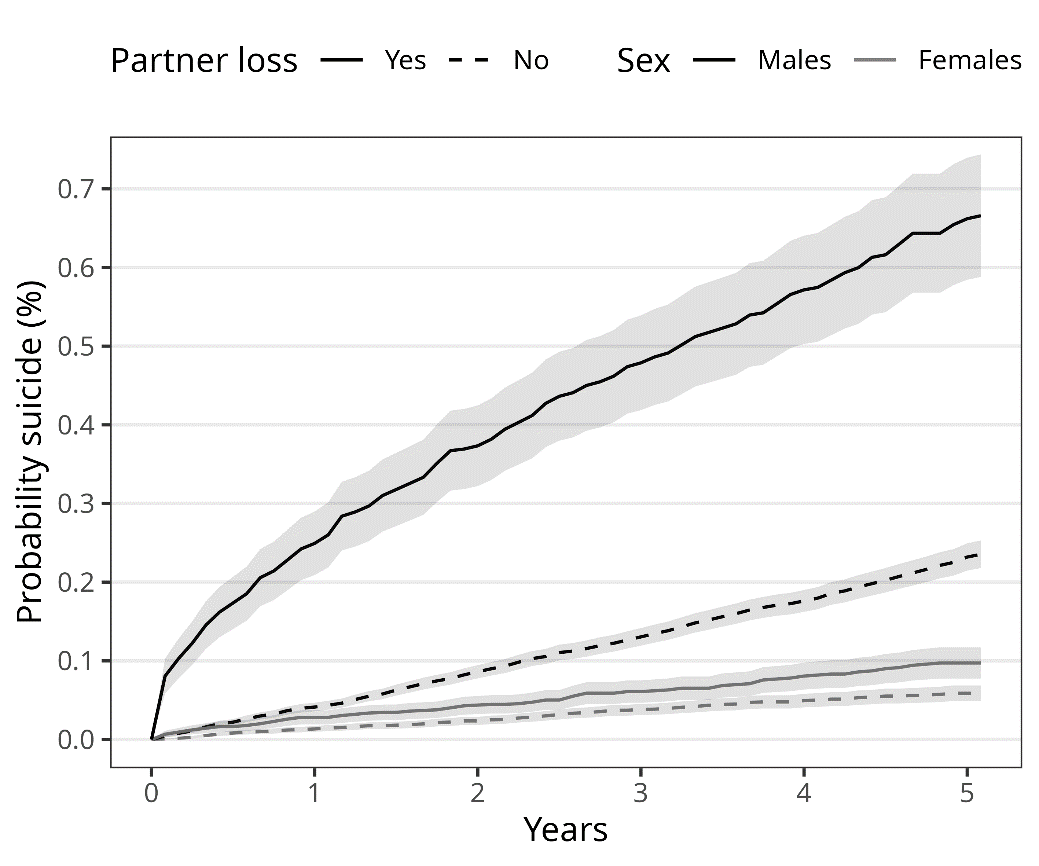
